# Machine-actionable criteria chart the symptom space of mental disorders

**DOI:** 10.1101/2025.09.12.25335630

**Authors:** Barbara Strasser-Kirchweger, Raoul Hugo Kutil, Georg Zimmermann, Christian Borgelt, Wolfgang Trutschnig, Florian Hutzler

**Affiliations:** Department of Psychology, Centre for Cognitive Neuroscience, University of Salzburg, Hellbrunnerstraße 34, Salzburg, 5020, Salzburg, Austria; Department Artificial Intelligence & Human Interfaces (AIHI), IDA Lab Salzburg, University of Salzburg, Jakob-Haringer-Straße 6, Salzburg, 5020, Salzburg, Austria; Team Biostatistics and Big Medical Data, Paracelsus Medical University, Strubergasse 21, Salzburg, 5020, Salzburg, Austria

**Keywords:** Disorder Delineation, Disorder Similarity, Machine-Actionable Representation, Differential Diagnosis, DSM-5, Computational Psychiatry, Diagnostic Overlap

## Abstract

Diagnostic manuals encode community consensus in prose yet offer no direct means to computationally evaluate the conceptual integrity of disorder definitions. We introduce a machine-actionable framework that translates narrative diagnostic criteria into the full symptom space—the exhaustive set of symptom combinations valid for each disorder. This approach enables charting of how these symptom spaces intersect, diverge, or subsume one another. Applied to representative DSM-5 disorders and to the emerging definition of Long COVID, the framework confirms clear boundaries among established disorders while highlighting substantial conceptual redundancy between Long COVID and mood or anxiety disorders. Whereas probabilistic models infer patterns from broad textual corpora, the proposed framework directly interrogates explicit consensus criteria, providing a transparent and reproducible means of assessing conceptual coherence. By making consensus-based diagnostic knowledge computable, the framework supports the refinement of classification systems and provides a foundation for interpretable clinical decision support.

## Introduction

Accurate diagnosis depends on clear, consistent rules—but overlapping symptoms and heterogeneous criteria across disorders often make the process complex and error-prone ^1,2^. Meanwhile, while AI tools are increasingly used to support clinical decisions, they often lack direct access to the structured, consensus-based knowledge embedded in diagnostic manuals; moreover, in mental health applications the lack of understandability — a combination of transparency and interpretability — undermines trust and limits clinical usefulness, especially when AI outputs cannot be matched to clinicians’ domain knowledge ^3^.

Diagnostic manuals such as the Diagnostic and Statistical Manual of Mental Disorders, Fifth Edition (DSM-5), encode this knowledge in explicit, rule-based criteria that define which combinations of symptoms warrant each diagnosis. Unlike probabilistic or large-language-model systems — which infer patterns from vast text corpora ^4^—these manuals embody a normative, community-endorsed consensus formed through structured expert deliberation and formal review, a process that confers institutional and clinical authority ^5,6^.

Critically, these consensus criteria are not merely descriptive. In DSM-5, each criterion gives rise to a set of valid symptom constellations—what we term *criteria-satisfying symptom combinations* (CSSCs). Clinicians must mentally juggle these CSSCs across multiple disorders to avoid misdiagnosis, a task that is combinatorially explosive and beyond unaided human cognition.

One prime example is Major Depressive Disorder (MDD), which is defined in the DSM-5 by means of five diagnostic criteria. The first of these specifies that at least five out of nine symptoms must be present, including either “depressed mood” or “loss of interest or pleasure” as a mandatory component. Notably, each of these “symptoms” is itself a symptom cluster that requires further differentiation into discrete elements, as detailed below. Even this single criterion yields over seven million valid CSSCs. Further complicating matters, commonly endorsed symptoms such as fatigue and sleep disturbances recur across the definitions of multiple disorders, including anxiety disorders, bipolar disorder, and certain personality disorders ^7,8^. This very overlap often produces similar symptom combinations, complicating differential diagnosis—that is, the task of distinguishing one disorder from others with possibly overlapping presentations. Accurate diagnosis remains essential, as misdiagnosis can lead to ineffective treatment, prolonged suffering, and significantly increased healthcare costs ^9^.

Although recent revisions to diagnostic manuals — such as the DSM-5 and ICD-11 — have enhanced specificity and diagnostic accuracy ^10,11^, their content remains largely narrative, limiting computational access and systematic refinement. Ressources like ICD–SNOMED (Systematized Nomenclature of Medicine) mappings have standardized terminologies^12,13,14^; SNOMED CT offers rich clinical hierarchies and relationships that facilitate encoding and inference in decision-support systems but does not itself encode the structured diagnostic rules found in manuals. Similarly, the Human Disease Ontology^15,16^ provides a formal, literature-informed disease *classification* in OWL—ideal for semantic interoperability and data integration—but it does not represent the specific symptom-combination logic underpinning DSM- or ICD-style criteria.

A standardized, machine-actionable representation of diagnostic rules would allow for the systematic identification and formal analysis of CSSCs and their potential overlap across disorders. Such a framework could support clinicians in navigating the full diagnostic space, highlight definitional inconsistencies, and facilitate the refinement of diagnostic manuals. To our knowledge, no existing system comprehensively encodes the logical relationships between criteria and symptoms as defined in community-accepted diagnostic manuals.

The need for such a system is particularly urgent in the context of emerging conditions. Long COVID, for example, remains poorly defined, with proposed diagnostic criteria differing across guidelines ^17,18,19,20,21^. Symptoms such as fatigue and cognitive dysfunction overlap significantly with those of established disorders, raising the question of whether proposed definitions produce CSSCs that are already fully covered by existing categories. Without a formal method to assess such overlap, emerging diagnostic constructs risk conceptual redundancy.

Aiming to overcome these problems, in this paper we present a machine-actionable framework for representing consensus-based diagnostic knowledge that enables the generation and analysis of all valid CSSCs. Using the DSM-5 as an illustrative example, we demonstrate how our framework enables analyses such as assessing diagnostic similarity and conceptual delineation between established disorders and emerging conditions like Long COVID—among other potential clinical and computational applications. We show how our approach identifies symptom combination overlap, quantifies conceptual delineation, and highlights definitional ambiguities—laying the groundwork for more rigorous, systematic, and scalable diagnostic developments.

In addition to supporting diagnostic knowledge refinement, our framework can also serve as the foundation for clinical recommendation systems that assist practitioners in the context of navigating complex symptom combinations and improving diagnostic accuracy in real time. As the integration of AI into clinical practice accelerates, such a framework provides the structured, consensus-based knowledge that is essential for trustworthy, explainable, and clinically valid AI systems.

## Main

### From narrative to formal representation

In order to create a machine-actionable representation of diagnostic knowledge, the descriptive language used in diagnostic manuals must be translated into formal representations. Specifically, symptoms and diagnostic criteria described in prose—for instance, in the DSM-5—must be transformed into logical structures specifying which symptoms are required or irrelevant for each disorder.

To illustrate, Fig. 1 (left panels) presents excerpts of the narrative diagnostic criteria for Major Depressive Disorder and Persistent Depressive Disorder, which define conditions under which specific symptom combinations are sufficient for diagnosis. These criteria are formalized into logical statements in Fig. 1 (upper right panel). For example, the requirement in Persistent Depressive Disorder that “two (or more)” of six symptoms be present is expressed as a cardinality constraint (e.g., “≥ 2”) in our formal language.

**Fig. 1:**
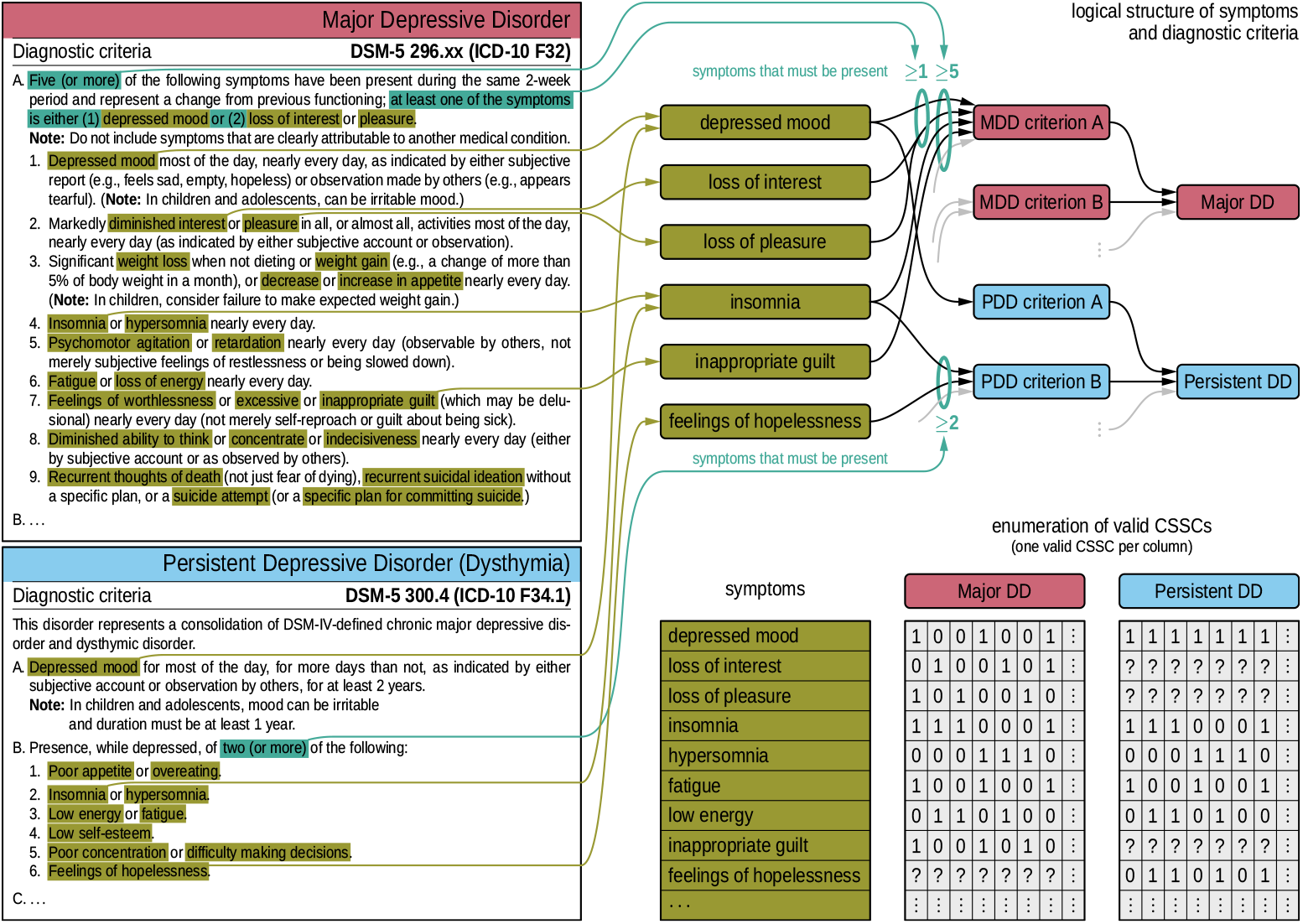
Left panels: Narrative diagnostic criteria provided in diagnostic manuals, such as DSM-5, are analyzed to identify key elements. Exemplary criteria are shown for Major Depressive Disorder (MDD,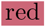) and Persistent Depressive Disorder (PDD, 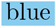). The analysis involves extracting essential components, such as specific symptom counts (e.g., “five or more”, highlighted in 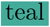) and individual symptoms (e.g., “depressed mood” or “loss of interest”, highlighted in 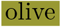). **Upper right panel:** The afore-mentioned elements are formalized within a structured framework, where symptoms, criteria, and disorders are depicted as distinct nodes, and numerical thresholds are represented as edges connecting specific symptoms to diagnostic criteria. This framework organizes the relationships between symptoms, diagnostic criteria, and disorders. Edges illustrate the rules that define these relationships, such as how a symptom connects to a specific criterion and how that criterion links to the broader disorder. For instance, when two disorders share a symptom, the same symptom node will have rules linking it to different criterion nodes for each disorder. This structured representation enables a clear depiction of overlapping criteria and symptoms across disorders, facilitating a systematic analysis of their interrelationships. **Lower right panel:** Binary encoding of the CSSCs of the two disorders. For MDD the vector in the first column represents a CSSC: the vector indicates symptoms that are present (1), those that are absent (0), and those that are irrelevant (“?”).

Importantly, symptoms not explicitly required by a disorder’s diagnostic criteria should not be interpreted as requiring absence. Their presence or absence carries no diagnostic value unless explicitly stated. For instance, “feelings of hopelessness” is not listed among the diagnostic criteria for Major Depressive Disorder. A patient may or may not report this symptom, but it does not affect the diagnosis. In our framework, such symptoms are considered *irrelevant* for that disorder (for details on formal definitions of relevance and redundancy, see Methods).

These logical structures, which constitute the intensional definition of a disorder, enable the systematic generation of all valid (CSSCs). CSSCs can be represented as binary vectors, with each dimension corresponding to a symptom: a value of 1 indicates presence, 0 indicates absence, and “?” denotes a diagnostically irrelevant symptom (see Fig. 1, lower right panel). To maintain binary integrity, irrelevant symptoms are implicitly treated as 0 during analysis (see Methods for details).

To further illustrate, consider a toy model with four possible symptoms: a, b, c, and d (see Fig. 2). Suppose that Disorder W requires (A) Symptom a and (B) at least one of b or c (Panel a). This yields three valid CSSCs: (1, 1, 1, ?), (1, 1, 0, ?), and (1, 0, 1, ?), where Symptom d is irrelevant and thus marked with “?” (Panel b). Disorder X, in contrast, requires (A) Symptom d and (B) at least one of b or c, resulting in (?, 1, 1, 1), (?, 1, 0, 1), and (?, 0, 1, 1), with Symptom a being irrelevant.

**Fig. 2:**
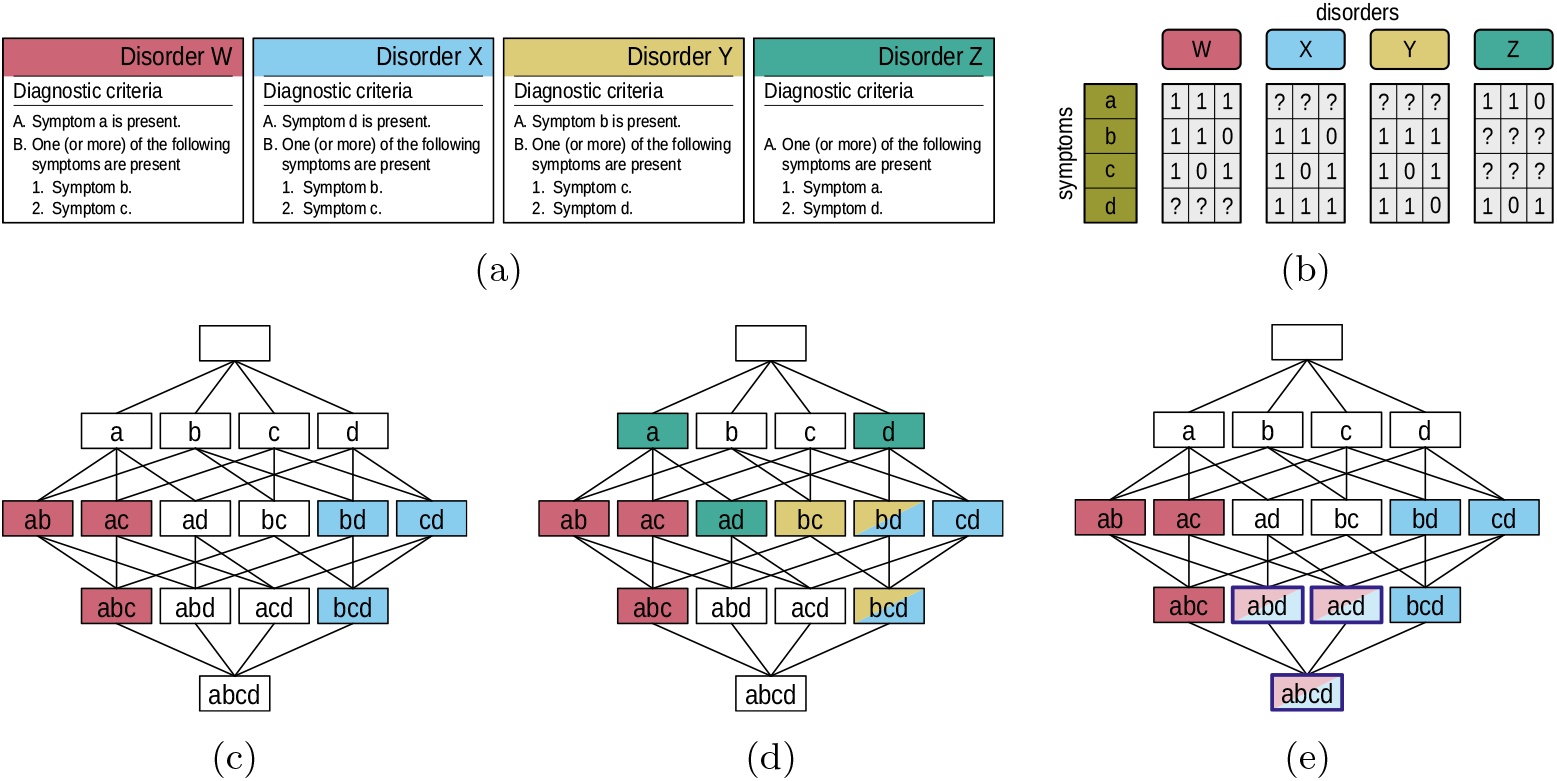
**(a)** Narrative descriptions of diagnostic rules for four hypothetical disorders: W, X, Y, and Z. **(b)** Binary vector encodings of valid criteria-satisfying symptom combinations (CSSCs) for each disorder. Columns are grouped by disorder; rows correspond to symptoms. Values of 1 and 0 denote relevant symptoms that are present or absent, respectively. Irrelevant symptoms are indicated by “?”. **(c)** The full fourdimensional symptom space visualized as a Hasse diagram, showing all possible symptom combinations. CSSCs for Disorders W 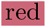 and X 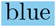 are overlaid. Irrelevant symptoms are implicitly treated as 0 (see Methods for details). **(d)** Same symptom space, now including CSSCs for Disorder Y 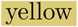 and Disorder Z 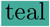. Weakly irredundant CSSCs (bd and bcd) shared by Disorders X 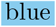 and Y 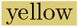 violate the *no-overlap requirement* (absence of identical CSSCs), indicating that the disorders are not conceptually separable, as they share at least one identical CSSC. This constitutes a violation of the *no-overlap requirement* and confirms the lack of diagnostic distinctiveness between Disorders X and Y. Symptoms a and d, representing CSSCs of Disorder Z 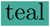, do not overlap with CSSCs of other disorders but are *subsumed* by CSSCs of W (ab, ac,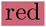) and X (bd, cd,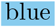), respectively—illustrating violations of the *no-subsumption requirement*. **(e)** Illustration of real-world patient presentations for Disorders W and X. Solid-colored nodes represent presentations corresponding with CSSCs, lighter nodes with dark outlines reflect presentations that include additional, non-required disorder-wise (diagnostically) irrelevant symptoms.

All possible combinations of these four symptoms define a four-dimensional symptom space, with each symptom corresponding to one dimension. Panel (c) uses a Hasse diagram to visualize this space, ranging from the empty set (top node) to the full set of symptoms (bottom node labeled “abcd”). Within this space, each disorder defines a subset corresponding to its valid CSSCs. This is illustrated for the subsets of Disorders W (red) and X (blue), overlaid on the full symptom space. Irrelevant symptoms (e.g., d for Disorder W, a for Disorder X) play no role in determining diagnostic membership and are implicitly treated as 0 (see Methods for details) when calculating, e.g., the degree of similarity of two disorders.

### Delineation of disorders

In Fig. 2, Panel (b), the nodes representing Disorders W and X correspond to *weakly irredundant* CSSCs — symptom combinations that exclude diagnostically irrelevant symptoms (see Methods for formal definition). These CSSCs form the basis for evaluating whether two disorders can be conceptually delineated—that is, whether their diagnostic criteria are sufficiently distinct to support reliable differential diagnosis.

To formalize this evaluation, we introduce two delineation requirements. The first is the *absence of identical CSSCs (in the following:* no-overlap requirement): two disorders should not share any weakly irredundant CSSCs. If such a CSSC were assigned to both disorders, it would be impossible to uniquely determine the diagnosis from that combination, thereby rendering differential diagnosis conceptually impossible. Such a case would reflect a structural flaw in the definitions themselves, as a single symptom profile would simultaneously fulfill two disorder definitions.

In the case of Disorders W and X, see Fig. 2, Panel (c), each CSSC node in the Hasse diagram is assigned to at most one disorder—either red for W or blue for X, but never both. Although the disorders share Symptoms b and c, they differ in requiring either a (for W) or d (for X), implying that no weakly irredundant CSSC is shared. Thus, disorders W and X satisfy the *no-overlap requirement*. To illustrate a failure of the *no-overlap requirement*, consider Disorder X (blue) and Y (yellow) presented in Fig. 2, Panel (d). X and Y share two weakly irredundant CSSCs: bd and bcd, indicating that differential diagnosis is logically impossible based on the current definitions.

The second delineation requirement is the *absence of subsumption (in the following:* no-subsumption requirement): no weakly irredundant CSSC of one disorder may be a 7 strict subset of any CSSC of another disorder (see Methods for a formal definition). Even if two disorders do not share identical CSSCs, delineation fails if one disorder’s CSSC is fully contained within another disorder’s CSSC. In such cases, the additional symptoms offer no meaningful distinction, and diagnostic boundaries collapse. This requirement is asymmetric and must be assessed from the perspective of each disorder individually.

As shown in Fig. 2, Panel (c), no weakly irredundant CSSC for Disorder W is a strict subset of any for Disorder X, and vice versa. This is visually represented in the Hasse diagram: no node of one disorder lies directly beneath a node of the other, connected via an edge. Accordingly, Disorders W and X also fulfill the *no-subsumption requirement*. Panel (d) illustrates a violation of the *no-subsumption requirement* for Disorder Z in green with respect to W and X. In fact, we can find weakly irredundant CSSCs of Z, namely a and d, which (while not being CSSCs of W and X) are strict subsets of CSSCs of W and X. Due to the asymmetry of the *no-subsumption requirement*, this subset relationship compromises delineation even if the reverse does not hold.

When conceptual delineation is not achievable, the unavoidable resulting diagnostic ambiguity calls into question the validity and coherence of the disorder definitions. Such cases suggest the need for conceptual refinement or the identification of additional, as-yet-undefined symptoms enabling reliable diagnostic separation.

It is also important to distinguish failures in conceptual delineation from the variability observed in real-world patients. Patients may exhibit additional symptoms beyond those required for diagnosis, or may meet criteria for multiple disorders when different sets of required symptoms are present. This distinction is illustrated in Fig. 2, Panel (e): a patient presenting with all four symptoms (a, b, c, and d) would meet the criteria for both Disorder W and Disorder X (they would, in fact, meet the criteria for all four disorders). Since each disorder treats one of those symptoms as irrelevant, both diagnoses would be valid and reflect comorbidity — not a failure of conceptual delineation.

To summarize, the approach outlined so far transforms narrative diagnostic criteria into a formal, machine-actionable framework enabling the systematic generation of all valid CSSCs for each disorder represented in the system. These combinations constitute a disorder’s derived combinatorial structure, providing a basis for rigorously evaluating the internal logic and conceptual coherence of diagnostic definitions, and for assessing whether two disorders are formally delineable (based on the absence of shared or subsumed CSSCs). Having access to the complete set of CSSCs for two disorders also enables quantitative comparisons, allowing us to move beyond binary delineability and measure the degree of similarity between disorders based on their weakly irredundant CSSCs, encoded as binary vectors.

### Quantifying conceptual relatedness

To quantify the degree of similarity between two disorders, we compute the Maximum Pairwise Cosine Similarity (MPCS) between their respective weakly irredundant CSSCs. Cosine similarity yields a similarity score between 0 and 1, where 1 indicates identity and 0 indicates no shared symptom presence. While a full mathematical description of the procedure is provided in Methods, the key steps are as follows: Given two disorders, X and Y, characterized by their sets of weakly irredundant CSSCs, we: (i) compute, for each CSSC in X, its maximum cosine similarity to any CSSCs in Y; (ii) aggregate these similarity values by either taking the mean or the maximum over the CSSCs of X; (iii) repeat the same procedure in reverse—from Y to X; (iv) take the maximum of the two resulting values, i.e., of the two maxima or the two means, yielding either MPCS_max_ or MPCS_mean_. Note that in order to compute cosine similarity, each CSSC must be embedded in a shared symptom space. This requires augmenting the vectors with the irrelevant symptoms from the other disorder (i.e., symptoms that are relevant for one disorder but irrelevant for the other are encoded as 0 for that disorder), ensuring a consistent dimensionality across comparisons. A value of MPCS_max_ = 1 indicates the presence of at least one identical CSSC between X and Y, violating the *no-overlap requirement*. Conversely, a value of 0 implies that the sets of relevant symptoms are completely disjoint — only in this case can both MPCS_max_ and MPCS_mean_ be zero. In all other cases, the presence of shared symptoms will necessarily produce nonzero similarity values, even when disorders are otherwise delineable (see Fig. 1).

Turning to our toy example, MPCS_mean_ values are 0.556 (W, X), 0.661 (W, Y), 0.939 (X, Y), 0.664 (W, Z), 0.664 (X, Z), and 0.426 (Y, Z). Disorders X and Y exhibit the highest *average* similarity, consistent with their shared CSSCs in Fig. 2 (see also Table 3).

To validate the proposed similarity measure, we applied our approach to a selected set of disorders within the schizophrenia spectrum—specifically, Delusional Disorder, Schizophreniform Disorder, Schizophrenia, and Schizoaffective Disorder — as defined by the DSM-5. To provide contrast, we included Speech Sound Disorder as a conceptually unrelated control. For each disorder, we enumerated the complete set of valid CSSCs. These were encoded as binary vectors, with dimensionality determined by the total number of unique symptoms involved.

Fig. 1 presents the MPCS_mean_ values for the selected disorders and confirms the expected conceptual separation of Speech Sound Disorder, which exhibited zero similarity with all schizophrenia spectrum disorders. This result serves as a plausibility check for our framework: a similarity score of zero implies that the sets of symptoms relevant for diagnosing the two disorders are entirely disjoint.

By contrast, the highest similarity was observed between Schizophrenia and Schizophreniform Disorder (MPCS_mean_ = 0.744), consistent with their close clinical and diagnostic relationship. Both disorders share nearly identical core symptom requirements — such as delusions, hallucinations, and disorganized speech — with the primary distinction being the duration of symptoms required for diagnosis ^7^. As a result, many CSSCs for one disorder closely resemble those of the other. This structural similarity reflects their clinical proximity as discussed below.

Building on these findings, the next section assesses whether such conceptually related disorders — notably Schizophrenia and Schizophreniform Disorder — not only exhibit high structural similarity but also fulfill the formal requirements for conceptual delineation.

### Delineation of established disorders

Building on our previous finding that Schizophrenia and Schizophreniform Disorder exhibit high structural similarity in their CSSCs, we now evaluate whether these two disorders fulfill the delineation requirements defined in Delineation of disorders. While their shared core symptom requirements account for the observed similarity, a failure to meet the delineation criteria would indicate a conceptual redundancy, precluding clear differential diagnosis.

As before, the analysis is based on the fully enumerated sets of weakly irredundant CSSCs for both disorders. To test the *no-overlap requirement* — the absence of identical CSSCs — we again applied the MPCS. Specifically, we computed MPCS_max_ values to identify whether any pair of CSSCs — one from each disorder — were fully identical (see Methods for details).

For the hypothetical disorders introduced in Fig. 2, MPCS_max_ values were: (W, X) = 0.667, (W, Y) = 0.816, and (W, Z), (X, Z), (Y, Z) = 0.707. The pair (X, Y) yielded a value of 1.000, indicating a failure of the *no-overlap requirement* due to at least one identical CSSC — consistent with the overlap depicted in Fig. 2.

In contrast, for Schizophrenia and Schizophreniform Disorder, MPCS_max_ = 0.800, confirming that no identical CSSCs exist between the two disorders. The *no-overlap requirement* is therefore satisfied.

To test the *no-subsumption requirement* — the absence of subsumption — we evaluated whether any weakly irredundant CSSC from one disorder was a strict subset of a CSSC from the other. Because subsumption is not symmetric, both directions were assessed separately. The analysis revealed that no CSSC from Schizophrenia was a subset of any from Schizophreniform Disorder, and vice versa. Thus, the *no-subsumption requirement* is also fulfilled.

Figure 3 visualizes the relationship between CSSCs of the two disorders using a Sammon projection, which embeds high-dimensional vectors — such as binary vectors in our case—into two dimensions while preserving pairwise distances (see Methods).

**Fig. 3:**
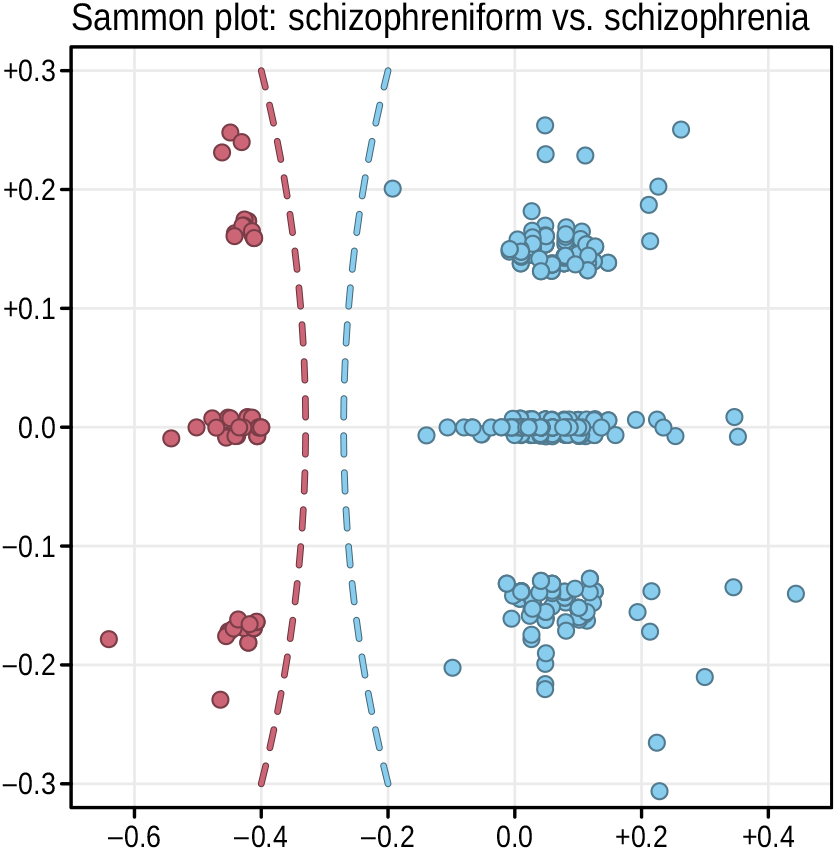
Sammon projection of the high-dimensional CSSCs for Schizophrenia 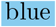 and Schizophreniform Disorder 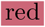, mapped into a two-dimensional space while preserving pairwise distances. The clear spatial separation between the two sets of binary vectors supports their conceptual delineation and illustrates that no identical CSSCs are shared between the disorders.

The resulting projection illustrates a clear separation between the combination sets, demonstrating fulfillment of the non-overlap criterion.

Finally, we examined which features accounted for delineation despite substantial symptom overlap. The key differentiating factor was symptom duration: Schizophrenia requires symptoms to persist for at least six months, whereas Schizophreniform Disorder is defined by a duration between one and six months. Schizophreniform Disorder is often diagnosed provisionally and may transition into Schizophrenia if symptoms persist beyond six months, a continuity that is well-documented both conceptually and epidemiologically ^7,22,23,24^.

While such an analysis remains tractable for pairwise comparisons, the computational complexity increases substantially when evaluating delineation across multiple disorders simultaneously — a challenge we revisit in the Discussion.

Our findings confirm that Schizophrenia and Schizophreniform Disorder satisfy both delineation requirements: they share neither identical nor subsumed CSSCs. This ensures that differential diagnosis is possible based solely on their formalized criteria. More broadly, this example illustrates how our framework enables the systematic evaluation of diagnostic coherence by leveraging the combinatorial structure of disorder definitions.

### Newly emerging clinical conditions

Having demonstrated how our approach can quantify similarity and assess delineation among established disorders, we now apply it to a newly emerging clinical condition: Long COVID. Characterized by persistent and often debilitating symptoms following acute SARS-CoV-2 infection, Long COVID currently lacks community-established, iteratively refined diagnostic criteria.

A recent consensus report by the U.S. National Academies of Sciences, Engineering, and Medicine (NASEM) defines Long COVID as a chronic, systemic disease state persisting for at least three months and involving one or more organ systems ^25^. Common symptoms include fatigue, cognitive dysfunction, and mood disturbances ^26^—features also observed in established disorders such as MDD, raising the potential for diagnostic ambiguity. The NASEM report emphasizes that “no published, standardized guidelines for developing disease definitions” currently exist for Long COVID ^25^. Instead, the proposed definition synthesizes multiple sources and emphasizes attribution, time course, clinical features, and exclusions. While appropriate for an evolving condition, this approach underscores the need for formal tools to evaluate whether emerging disorder definitions are conceptually distinct.

Long COVID presents a compelling test case for our framework. The definition proposed by Ely et al. ^26^ organizes Long COVID into three conceptual levels. Level 1 specifies symptoms or symptom clusters commonly reported in affected individuals, such as persistent fatigue, post-exertional malaise, and cognitive impairments like difficulty concentrating. These symptoms form the definitional core from which candidate diagnostic criteria can be constructed. Level 2 addresses co-occurring diagnosable conditions—including cardiovascular disease, arrhythmias, and mood disorders—which are treated either as differential diagnoses or comorbidities. This level provides a natural comparison point: we assess whether the symptom combinations implied by Long COVID are formally delineable from those of established disorders in this set. Level 3 captures broader contextual features of the condition, including diagnostic setting, applicability across age groups, and its relapsing–remitting or progressive course. While clinically relevant, these features are not formally encoded in our combinatorial framework and thus fall outside the scope of the present analysis.

The central question is whether the symptom combinations implied by Long COVID’s Level 1 definition give rise to a set of CSSCs that are formally delineable from those of established disorders, particularly those listed as differential diagnoses in Level 2.

To evaluate the *no-overlap requirement* (i.e., the absence of identical criteria-satisfying combinations), we compared the set of CSSCs derived from Level 1 Long COVID definition to those of nine established DSM-5 disorders spanning neurodevelopmental, psychotic, depressive, and anxiety-related categories (see Symptom harmonization for details). Following the procedure introduced earlier, we computed the Maximum Pairwise Cosine Similarity (MPCS) for each disorder pair. Specifically, we used MPCS_max_ to identify whether there is at least one identical combination. Table 2, column MPCS_max_, summarizes the results.

Long COVID showed no similarity to schizophrenia spectrum disorders, Delusional Disorder, or Speech Sound Disorder, all yielding MPCS_max_ values of 0. Substantial similarity values were observed with depressive and anxiety-related disorders, including Persistent Depressive Disorder (0.816), Panic Disorder (0.816), Major Depressive Disorder (0.756), and Generalized Anxiety Disorder (0.707). Despite substantial similarity with mood and anxiety disorders, no disorder in our comparison set shared any identical CSSC with Long COVID — that is, no MPCS_max_ reached 1. Thus, the *no-overlap requirement* is satisfied.

To evaluate the *no-subsumption requirement*, we assessed whether any weakly irredundant CSSC of an established disorder is criteria satisfying for Long COVID, or vice versa. No subsumption was observed in either direction for Speech Sound Disorder, Delusional Disorder, or any of the schizophrenia spectrum disorders, confirming their conceptual separation from Long COVID.

In contrast, several mood and anxiety disorders violated this requirement. A small number of Long COVID combinations were strict subsets of valid CSSCs from Major Depressive Disorder (*N*_sub_ = 15), Persistent Depressive Disorder (15), Panic Disorder (15), and Generalized Anxiety Disorder (7). Conversely, over 1.29 billion of the 1.38 billion valid CSSCs for Major Depressive Disorder (94%) satisfied the Level 1 criteria for Long COVID. Similar proportions were observed for the other mood and anxiety disorders, indicating substantial conceptual overlap (see Table 2).

These findings carry significant implications for both clinical practice and research. The fact that over 90% of valid symptom combinations for major depressive and anxiety disorders satisfy the current Level-1 definition of Long COVID suggests a diagnostic framework that is too permissive to ensure conceptual distinctiveness. Without additional constraints — such as symptom chronology, exclusion logic, or biomarkers — this broad definition may overestimate prevalence and blur distinctions between clinical entities, reducing diagnostic precision in treatment and surveillance. In this light, formal delineation testing offers more than conceptual clarity: it becomes an essential safeguard against diagnostic inflation and category collapse.

We may be witnessing a case of *diagnostic drift*: although adding a history of COVID-19 infection could, in principle, restore delineation, this constraint has become practically meaningless. In a post-pandemic world, prior infection applies to the vast majority of the population and thus functions as a *trivial predicate* in the logical sense—true for (nearly) all. While such an addition might formally enforce delineation, it would do so at the expense of clinical utility.

## Discussion

### Formalizing and quantifying similarity

This work demonstrates a proof of principle for systematically transforming narrative diagnostic knowledge — as codified in consensus manuals such as the DSM-5 — into a formalized, machine-readable, and machine-actionable representation, which can then be further analyzed. By translating diagnostic criteria into logical structures and binary vectors we enabled direct access to the full extension of disorder definitions and established the means to evaluate their conceptual coherence.

We introduced two formal delineation requirements — the absence of identical CSSCs (*no-overlap requirement*) and the absence of subsumption (*no-subsumption requirement*) — as necessary conditions for distinguishing between disorder definitions in a logically coherent diagnostic framework. Applying these criteria, we first validated the framework using established disorders, showing that Schizophrenia and Schizophreniform Disorder, despite substantial overlap in symptom criteria, fulfill both delineation requirements and can be distinguished on the basis of duration. We then applied the same framework to Long COVID as an emerging clinical condition. While no identical combinations were identified, several CSSCs derived from its candidate definition were strictly subsumed by CSSCs of existing depressive and anxiety disorders.

This asymmetry compromises Long COVID’s conceptual independence and highlights the need for delineation testing in the development of new diagnostic categories. More broadly, our findings show how formal representations of diagnostic criteria can expose conceptual redundancies, inform refinement of disorder definitions, and support more rigorous, evidence-based expansion of diagnostic systems.

Going beyond determining whether disorders can be delineated, our approach allows to quantify the extent of similarity of pairs of disorders on a scale from 0 to 1. The higher the obtained score MPCS_mean_ the more similar the disorders are.

### Conceptual implications

The approach described here shows that a formal, machine-actionable representation of diagnostic knowledge can resolve core conceptual challenges in diagnostic classification. While narrative diagnostic criteria are interpretable by humans, they often obscure logical relationships between disorders, making it difficult to assess whether definitions are meaningfully distinct.

By encoding diagnostic rules into logical structures and generating the full space of criteria-satisfying symptom combinations (CSSCs), this framework reveals overlaps that may remain hidden in narrative form. That emerging definitions—such as that of Long COVID—can yield combinations fully subsumed under established disorders underscores the need for rigorous, formal tools to evaluate diagnostic coherence.

These findings also expose the scale of complexity clinicians must navigate. For example, Major Depressive Disorder yields billions of valid CSSCs. Manually comparing such enormous numbers is infeasible, particularly as the number of disorders and comparisons grows. Without formalized, machine-supported representations, ensuring conceptual delineation across a growing diagnostic landscape becomes practically impossible.

Crucially, the proposed framework also enables the iterative refinement of diagnostic manuals. As new conditions arise and criteria evolve, formal delineation testing at the level of symptom combinations offers a scalable mechanism for validating, whether a proposed definition is truly distinct — supporting clearer boundaries, improved differential diagnosis, and more coherent classification systems.

### Methodological insights

While the framework presented here provides a foundation for machine-actionable diagnostic reasoning, its implementation raises several technical and methodological challenges.

A core challenge is the translation of narrative diagnostic criteria into formal logic. Diagnostic manuals are written for clinical interpretability, not computational precision. As such, they often contain semantically ambiguous expressions—such as “weight change”, which implicitly covers both weight gain and weight loss—that are easily understood by clinicians but must be disambiguated explicitly for algorithmic processing. Effective translation therefore requires expert-guided modeling of symptom boundaries to ensure mutual exclusivity and logical consistency.

A second challenge lies in the computational scale of the problem. For disorders with multiple symptoms, the number of valid symptom combinations grows exponentially. As demonstrated in this study, even a single diagnostic criterion can yield millions of valid CSSCs. Enumerating and storing these combinations—especially across multiple disorders—places significant demands on memory, computation, and runtime, requiring the use of high-performance computing infrastructure and tailored optimization techniques.

Beyond practical concerns, these challenges are rooted in fundamental computational theory. Determining whether two disorders share any valid symptom combination can be reduced to a variant of the propositional satisfiability problem — a canonical NP-complete problem in computer science ^27,28^.

Taken together, these challenges emphasize that machine-actionable models of diagnostic knowledge are not only technically feasible, but also scientifically non-trivial. Their successful implementation depends on careful logical modeling, rigorous complexity-aware design, and close collaboration between clinical experts and computational scientists.

### Advantages over probabilistic models

A central distinction between the deterministic framework described in this study and current Machine Learning approaches lies in their relationship to diagnostic consensus. Large language models (LLMs), including those recently evaluated for clinical use ^4^, are trained on vast corpora of biomedical literature, web content, and other text sources to reproduce statistically frequent linguistic patterns. While this allows them to generate plausible-sounding outputs across a wide range of queries, these systems lack any internal representation of codified diagnostic criteria—and no mechanism to ensure that the symptom combinations they propose conform to community-endorsed standards.

By contrast, the deterministic system introduced here does not infer diagnostic logic from data; it encodes it explicitly. Diagnostic rules are formalized based on structured, consensus-driven criteria such as those found in the DSM-5, allowing the system to produce decisions only when warranted—and to abstain otherwise. This prevents the generation of diagnoses for which no formal justification exists. This is particularly crucial in clinical contexts, where explainability, consistency, reproducibility, and adherence to established consensus are essential.

Recent evaluations underscore the significance of this distinction. In one benchmark, the Med-PaLM model aligned with expert consensus in 79.5% of test cases ^4^. While promising, this figure highlights a fundamental limitation: probabilistic systems approximate consensus — they do not instantiate it.

In clinical diagnosis, where adherence to established standards is non-negotiable, even minor deviations from standard criteria can impact diagnostic decisions and reliability. Our system is designed to be fully transparent while explicitly endorsing community standards and consensus frameworks for clinical reasoning, thereby strengthening both interpretability for clinicians and alignment with established diagnostic practices.

Moreover, the consensus codified in diagnostic manuals is not a statistical artifact but a normative construct: it reflects structured expert deliberation, procedural transparency, and collective endorsement ^5,6^. LLMs, by design, conflate frequency with authority. They externalize the consensus process, creating the risk that regularities in language usage might be mistaken for explicit, community-governed standards. This approach may suffice for information retrieval, but it is less suited for domains like diagnosis, where rule-based clarity and reproducibility are central.

In this context, deterministic systems provide a necessary complement to probabilistic models. Rather than replacing consensus, this approach makes it computable—providing auditable and rule-based support grounded in formal clinical logic.

### Broader applications and future directions

The systematic representation and combinatorial analysis of diagnostic knowledge presented here create new possibilities, both for clinical practice and the development of a new field of diagnostic knowledge engineering. By making the full space of CSSCs accessible, this framework enables a level of diagnostic reasoning that exceeds the capacity of unaided clinical judgment—particularly when dealing with disorders characterized by vast combinatorial complexity. Disorders like Major Depressive Disorder can yield millions of valid symptom combinations, far beyond what clinicians can reliably track, especially under time pressure. In such contexts, heuristic reasoning—while efficient—may introduce biases that compromise diagnostic accuracy ^29,30^.

One immediate application is the development of evidence-based recommender systems for clinical use. They could systematically identify symptoms that are most discriminative for candidate diagnoses, guiding clinicians toward targeted, efficient, and transparent differential diagnosis. They would enable real-time comparison of patient presentations against the full set of CSSCs across all disorders — an analytic capability that has, until now, been impractical without computational assistance. Rather than replacing clinical expertise, this deterministic framework supports it by offering transparent, rule-based, reproducible decision-making build on the solid grounds of consensus knowledge.

Structured systems could also serve educational purposes, exposing users to nuanced differentiations between closely related disorders and thereby improving diagnostic training. Moreover, by reducing reliance on heuristic shortcuts under time pressure, these tools may help mitigate cognitive biases and improve consistency in clinical assessments.

Beyond individual diagnosis, this formalized framework offers a foundation for refining and updating diagnostic manuals themselves. Exhaustive comparisons between disorders could help expert committees evaluate the coherence of proposed criteria, identify unintended conceptual shortcomings, and clarify overlapping constructs. These capabilities also support the development of differentiated criteria early in the process of defining new disorders and promote alignment across related disorder domains.

Although this study focused on disorder definitions as encoded in narrative manuals, the framework could be extended to include diagnostic tools — such as structured interviews or symptom checklists — provided these tools are formally defined. This would enable not only the representation of disorder logic but also the efficient operationalization of next-step diagnostic procedures, guiding clinicians toward tools that best capture relevant symptoms based on a patient’s presentation.

Finally, as emerging conditions such as Long COVID illustrate, the ability to test whether a candidate disorder definition yields conceptually delineable symptom combinations is critical for ensuring that diagnostic categories remain clinically useful and scientifically valid. By codifying definitions formally and enabling precise, consensus-based reasoning, such systems reduce the risk of hallucination or normative drift — a gradual misalignment from formally accepted diagnostic standards. In this way, structured, machine-readable representations of diagnostic knowledge offer not only a path toward more reliable decision-making but also a platform for the continuous refinement of medical classification systems.

## Data Availability

Most of the data produced are available online at https://github.com/raoul-k/AIDA-Path/tree/main/data.
Some data in the present study are available upon reasonable request to the authors.

https://github.com/raoul-k/AIDA-Path

## Methods

### Definitions of core notions

Let *S* ={ *a, b, c, d*, … } be a fixed set of symptoms. A CSSC for a disorder is any subset *C* ⊆ *S* that fulfills the formal diagnostic criteria of that disorder. The criteria for each disorder, denoted X, Y, etc., can be defined by a logical rule over *S* that determines which subsets *C* qualify as CSSCs. The set of all CSSCs for a disorder X is referred to as its *extension*.

A CSSC *C* is called *irredundant* if no proper subset of *C* also qualifies as a CSSC. Otherwise, *C* is *redundant*. A symptom *s* ∈ *S* is considered *relevant* for disorder X if it appears in at least one irredundant CSSC of X. Otherwise, it is *irrelevant*. A CSSC is called *weakly irredundant* if it contains only relevant symptoms. It is called *strongly redundant* if it includes any irrelevant symptom. In our binary vector encodings, irrelevant symptoms are denoted by “?”, and are implicitly treated as absent during evaluation.

To assess the delineation of disorders, we define two requirements. The *no-overlap requirement* is violated if there exists at least one CSSC that is weakly irredundant for both X and Y. The *no-subsumption requirement* is violated if there exists at least one CSSC that is weakly irredundant for X and also satisfies the criteria of Y, even if not weakly irredundant for Y. In this case, disorder X is said to *subsume* disorder Y. Subsumption is asymmetric and strictly weaker than overlap.

Illustrative examples for each of these core notions, with references to figures and tables in the main text, are provided in the Supplementary Methods.

### Cosine similarity

To enable quantitative comparison of disorders, each criteria-satisfying symptom combination (CSSC) is represented as a binary vector over the shared symptom space, with 1 indicating presence, 0 absence of relevant symptoms, and “?” indicating irrelevant symptoms (implicitly treated as absent during evaluation and encoded as 0). Each disorder is thus described by a binary matrix *A* ∈ {0, 1 } ^*n×m*^, where columns correspond to CSSCs and rows to symptoms.

Similarity between two disorders is assessed via *pairwise cosine similarity*. For binary (or general) vectors *A* = (*A*_1_, …, *A*_*n*_) and *B* = (*B*_1_, …, *B*_*n*_), cosine similarity is the cosine of the angle between the two vectors and defined as

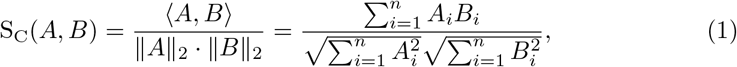

where ⟨·, ·⟩ denotes the inner product and ∥ · ∥ _2_ the Euclidean norm ^1^. For binary vectors *A, B* equation (1) reduces to

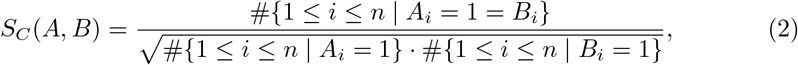

Here, # denotes set cardinality. While cosine similarity generally ranges from [− 1, 1], it is restricted to [0, 1] for binary vectors. To ensure comparability, all CSSCs are embedded in a shared symptom space of equal dimensionality.

Given two binary matrices **A** and **B** representing the full sets of CSSCs for disorders X and Y, respectively, we quantify their similarity using either the maximum or the mean of pairwise cosine similarities. For simplicity, let *A*_1_, …, *A*_*m*_ denote the columns of **A** and *B*_1_, …, *B*_*l*_ the columns of **B**.

In both approaches, we first compute the Maximum Pairwise Cosine Similarity (MPCS): for each fixed column *A*_*i*_ in **A**, we calculate its cosine similarity with all columns *B*_*j*_ in **B** and retain the maximum value. Mathematically, we compute:

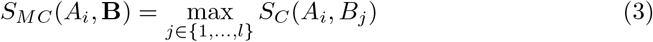

for every *i* ∈ {1, …, *m*. } These highest similarities *S*_MC_(*A*_1_, **B**), …, *S*_MC_(*A*_*m*_, **B**) are then aggregated, either by taking the maximum or the mean, depending on the purpose of the computation. Specifically, we define:

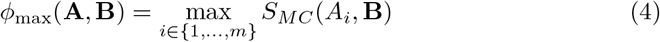

Or

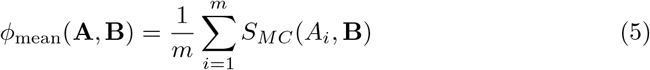

We refer to *ϕ*_max_ and *ϕ*_mean_ as maximum and mean similarity of **A** to **B**, respectively. Reversing the roles and repeating the process yields *ϕ*_max_(**B, A**) as well as *ϕ*_mean_(**B, A**). The final Maximum Pairwise Cosine Similarity is then either the quantity

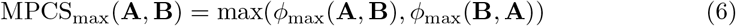

or the number

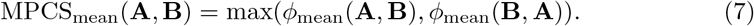

Obviously we always have

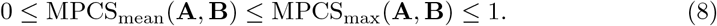

Applying the above-mentioned procedure to the toy example from Figure 2 yields the MPCS values in Table 3.

### Sammon mapping

Sammon mapping is a nonlinear dimensionality reduction technique that projects high-dimensional data into a lower-dimensional space while preserving pairwise distances as faithfully as possible ^2^. Unlike principal component analysis (PCA), which identifies linear components that maximize variance in the data, Sammon mapping prioritizes the preservation of pairwise distances, making it particularly suited for visualizing cluster structure in binary vector data such as CSSCs; in our setting, it outperformed both linear and nonlinear multidimensional scaling (MDS) in producing informative separations. The method minimizes Sammon’s stress function *E*, defined as:

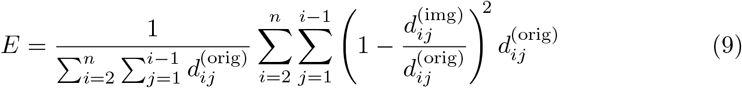

where 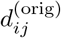 and 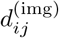 denote the distances between objects *i* and *j* in the original and image space, respectively. Larger original distances are weighted more heavily, emphasizing the preservation of broader structural relationships.

The optimization is typically initialized using PCA coordinates and refined via gradient descent. The normalized stress *E* quantifies how well interpoint distances are preserved: lower values indicate better fidelity, with *E* = 0 representing perfect preservation.

### Symptom harmonization

To support the formal analysis in Section 3, we harmonized the symptom terminology of Long COVID and comparator disorders to ensure representational consistency. Due to the string-based nature of our framework and the absence of standardized vocabulary terms at the symptom level, narrative descriptions from the 2024 NASEM report ^3^ were systematically aligned with DSM-5-compatible phrasing. All alignments are documented in the Supplementary Methods.

Diagnostic criteria for DSM-5 comparator disorders were further streamlined by removing elements that were unspecific or diagnostically non-differentiating. This ensured that comparisons reflected the definitional core of each disorder rather than general clinical disclaimers. Full mapping procedures and illustrative examples are also provided in the Supplement.

## Code Availability

The code used to generate the results in this paper is available in a public GitHub repository at:

https://github.com/raoul-k/AIDA-Path.

## Data Availability

The source data supporting the findings of this study, including the data used in Table 1 and Figure 3 and the associated calculated values, are available in a public GitHub repository at:

**Table 1:**
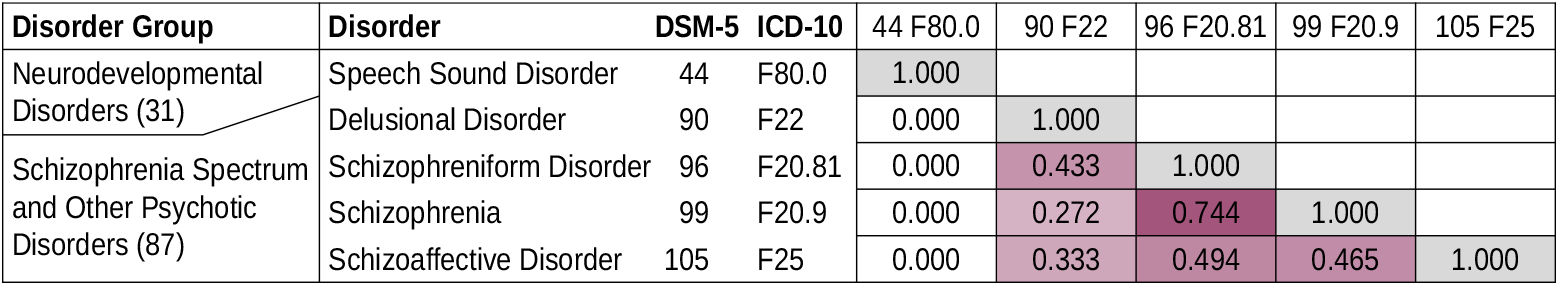
Obtained pairwise MPCS_mean_ values for the disorders mentioned in Quantifying conceptual relatedness. Zero similarity was observed between Speech Sound Disorder and each of the schizophrenia spectrum disorders, confirming their conceptual separation: A value of zero indicates that the sets of symptoms relevant for diagnosing one disorder are entirely disjoint from those of the other. By contrast, a gradient of similarity was observed within the schizophrenia spectrum, with the highest MPCS_mean_ value found between Schizophrenia and Schizophreniform Disorder.

**Table 2:**
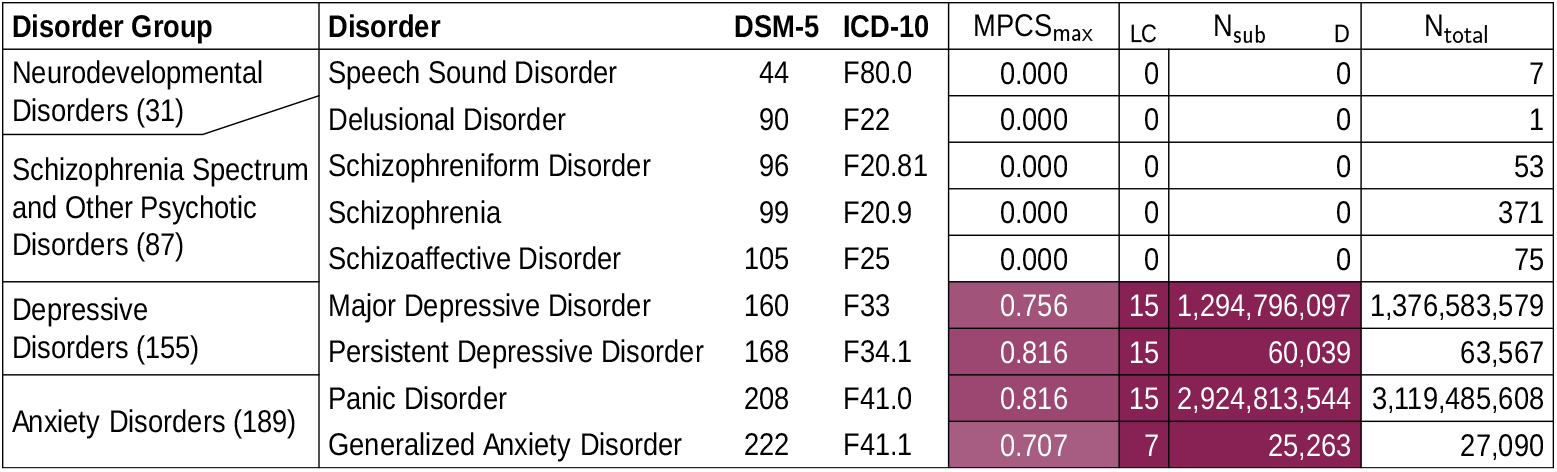
Comparison of criteria-satisfying symptom combinations (CSSCs) derived from the Level 1 definition of Long COVID with those of selected DSM-5 disorders to assess delineation requirements. The *no-overlap requirement* — absence of identical combinations, evaluated via the column MPCS_max_ — reports the maximum pairwise cosine similarity between any Long COVID CSSC and CSSCs of each comparator disorder. The no-overlap requirement is assessed via the MPCS_max_ column, where values below 1 indicate that no identical combinations were found. The *no-subsumption requirement* — absence of subsumption — is addressed in the subsequent columns. Column *N*_sub_ (LC) reports the number of weakly irredundant CSSCs of Long COVID that are strict subsets of at least one weakly irredundant CSSC of an established disorder and column *N*_sub_ (D) reports the number of weakly irredundant CSSC of an established disorder that are criteria-satisfying for Long COVID (i.e. are supersets of at least one weakly irredundant CSSCs of Long COVID). For reference, *N*_total_ provides the total number of weakly irredundant CSSCs for each established disorder.

**Table 3:** MPCS values for the toy example from Figure 2. The cell highlighted in red indicates a violation of the *no-overlap* requirement, as it reveals that at least one CSSC is valid for both X and Y—that is, the two disorders share an identical criteriasatisfying symptom combination. This result aligns with the overlapping node colors (yellow and blue) in the Hasse diagram shown in Figure 2 (d).

| MEAN | W | X | Y | Z |  | MAX | W | X | Y | Z |
| --- | --- | --- | --- | --- | --- | --- | --- | --- | --- | --- |
| W | 1.000 |  |  |  |  | W | 1.000 |  |  |  |
| X | 0.556 | 1.000 |  |  |  | X | 0.667 | 1.000 |  |  |
| Y | 0.661 | 0.939 | 1.000 |  |  | Y | 0.816 | 1.000 | 1.000 |  |
| Z | 0.664 | 0.664 | 0.428 | 1.000 |  | Z | 0.707 | 0.707 | 0.707 | 1.000 |

https://github.com/raoul-k/AIDA-Path/tree/main/data

## Supplementary Information

Supplementary information for Methods is available.

## Author Contributions

- **Conceptualization**: Barbara Strasser-Kirchweger, Florian Hutzler, Raoul Hugo Kutil, Christian Borgelt
- **Methodology**: Raoul Hugo Kutil, Christian Borgelt, Wolfgang Trutschnig
- **Formal Analysis**: Raoul Hugo Kutil, Christian Borgelt, Wolfgang Trutschnig
- **Writing – Original Draft**: Barbara Strasser-Kirchweger, Raoul Hugo Kutil, Florian Hutzler, Christian Borgelt
- **Writing – Review & Editing**: Florian Hutzler, Christian Borgelt, Raoul Hugo Kutil, Barbara Strasser-Kirchweger, Wolfgang Trutschnig, Georg Zimmermann
- **Supervision**: Florian Hutzler, Christian Borgelt
- **Validation**: Wolfgang Trutschnig, Georg Zimmermann
- **Visualization**: Christian Borgelt

## Main References

1. Singh, H., Schiff, G. D., Graber, M. L., Onakpoya, I. & Thompson, M. J. The global burden of diagnostic errors in primary care. BMJ Qual Saf 26, 484–494. ISSN: 2044-5415. eprint: https://qualitysafety.bmj.com/content/26/6/484.full.pdf. https://qualitysafety.bmj.com/content/26/6/484 (2017).

2. Ely, J. W., Graber, M. L. & Croskerry, P. Checklists to reduce diagnostic errors. Acad Med 86, 307–313. ISSN: 1040-2446. http://links.lww.com/ACADMED/A38 (Mar. 2011).

3. Joyce, D. W., Kormilitzin, A., Smith, K. A. & Cipriani, A. Explainable artificial intelligence for mental health through transparency and interpretability for understandability. npj Digital Medicine 6, 6 (2023).

4. Singhal, K. et al. Large language models encode clinical knowledge. Nature 620, 172–180 (2023).

5. Beatty, J. in Landscapes of Collectivity in the Life Sciences (eds Gissis, S. B., Lamm, E. & Shavit, A.) 179–198 (MIT Press, Cambridge, MA, 2017).

6. Wegwarth, O. & Gigerenzer, G. Torture by Consensus? On the Misuse of Evidence-Based Guidelines in Medicine. Medicine, Health Care and Philosophy 25, 329–338. https://doi.org/10.1007/s11019-022-10029-y (2022).

7. American Psychiatric Association. Diagnostic and statistical manual of mental disorders 5th. 10.1176/appi.books.9780890425596 (American Psychiatric Association, Washington, DC, 2013).

8. Newson, J. J., Pastukh, V. & Thiagarajan, T. C. Poor Separation of Clinical Symptom Profiles by DSM-5 Disorder Criteria. Front Psychiatry 12. ISSN: 1664-0640. https://www.frontiersin.org/journals/psychiatry/articles/10.3389/fpsyt.2021.775762 (2021).

9. Christensen, M. K. et al. The cost of mental disorders: a systematic review. Epidemiol Psychiatr Sci 29, e161. ISSN: 2045-7960 (Print) 2045-7960 (2020).

10. Horwitz, A. V. DSM: A History of Psychiatry’s Bible ISBN: 9781421440699 (Johns Hopkins University Press, Baltimore, 2021).

11. World Health Organization. International Classification of Diseases, 11th Revision (ICD-11) https://icd.who.int/browse11. xAccessed: 2025. 2019.

12. Cornet, R. & de Keizer, N. Forty years of SNOMED: a literature review. BMC Medical Informatics and Decision Making 8, 1–6 (2008).

13. Chang, E. & Mostafa, J. The use of SNOMED CT, 2013-2020: a literature review. Journal of the American Medical Informatics Association 28, 2017–2026 (2021).

14. Brooks, P. International Classification of Diseases, Version 10 - Clinical Modification — NCBO BioPortal May 2024. https://bioportal.bioontology.org/ontologies/ICD10CM.

15. Schriml, L. M. et al. Disease Ontology: a backbone for disease semantic integration. Nucleic acids research 40, D940–D946 (2012).

16. Schriml, L. M. et al. Human Disease Ontology 2018 update: classification, content and workflow expansion. Nucleic Acids Research 47, D955–D962 (2019).

17. Srikanth, S., Boulos, J. R., Dover, T., Boccuto, L. & Dean, D. Identification and diagnosis of long COVID-19: a scoping review. Progress in Biophysics and Molecular Biology 182, 1–7 (2023).

18. Sisó-Almirall, A. et al. Long Covid-19: proposed primary care clinical guidelines for diagnosis and disease management. International Journal of Environmental Research and Public Health 18, 4350 (2021).

19. Duerlund, L. S., Shakar, S., Nielsen, H. & Bodilsen, J. Positive predictive value of the ICD-10 diagnosis code for long-COVID. Clinical Epidemiology 14, 783–789 (2022).

20. Soriano, J. B., Murthy, S., Marshall, J. C., Relan, P. & Diaz, J. V. A clinical case definition of post-COVID-19 condition by a Delphi consensus. The Lancet Infectious Diseases 22, e102–e107 (2022).

21. Brown, D. A. & O’Brien, K. K. Conceptualising long COVID as an episodic health condition. BMJ Global Health 6, e007004 (2021).

22. Tandon, R., Nasrallah, H. A. & Keshavan, M. S. Schizophrenia, “just the facts” 4. Clinical features and conceptualization. Schizophrenia Research 110, 1–23 (2009).

23. Perälä, J. et al. Lifetime prevalence of psychotic and bipolar I disorders in a general population. Archives of General Psychiatry 64, 19–28 (2007).

24. Marneros, A. & Akiskal, H. S. The Overlap of Affective and Schizophrenic Spectra (Cambridge University Press, Cambridge, UK, 2007).

25. National Academies of Sciences, Engineering, and Medicine. A Long COVID Definition: A Chronic, Systemic Disease State with Profound Consequences (eds Fineberg, H. V., Brown, L., Worku, T. & Goldowitz, I.) ISBN: 978-0-309-71908-7. https://nap.nationalacademies.org/catalog/27768/a-long-covid-definition-a-chronic-systemic-disease-state-with (The National Academies Press, Washington, DC, 2024).

26. Ely, E. W., Brown, L. M. & Fineberg, H. V. Long Covid Defined 2024. eprint: https://www.nejm.org/doi/pdf/10.1056/NEJMsb2408466. https://www.nejm.org/doi/full/10.1056/NEJMsb2408466.

27. Cook, S. A. The Complexity of Theorem-Proving Procedures in Proceedings of the Third Annual ACM Symposium on Theory of Computing (STOC ‘71) (ACM, New York, NY, USA, 1971), 151–158.

28. Karp, R. M. in Complexity of Computer Computations (eds Miller, R. E. & Thatcher, J. W.) 85–103 (Springer, Boston, MA, 1972).

29. Korteling, J. H., van de Boer-Visschedijk, G. C., Blankendaal, R. A., Boonekamp, R. C. & Eikelboom, A. R. Human-versus artificial intelligence. Frontiers in Artificial Intelligence 4, 622364 (2021).

30. Klein, J. G. Five pitfalls in decisions about diagnosis and prescribing. BMJ: British Medical Journal 330, 781–783 (2005).

## Methods References

1. Salton, G. & McGill, M. J. Introduction to Modern Information Retrieval ISBN: 9780070544840 (McGraw-Hill, New York, 1983).

2. Sammon, J. A Nonlinear Mapping for Data Structure Analysis. IEEE Transactions on Computers C-18, 401–409 (1969).

3. National Academies of Sciences, Engineering, and Medicine. A Long COVID Definition: A Chronic, Systemic Disease State with Profound Consequences (eds Fineberg, H. V., Brown, L., Worku, T. & Goldowitz, I.) ISBN: 978-0-309-71908-7. https://nap.nationalacademies.org/catalog/27768/a-long-covid-definition-a-chronic-systemic-disease-state-with (The National Academies Press, Washington, DC, 2024).

