## Supplementary Information for "Machine-actionable criteria chart the symptom space of mental disorders"

September 12, 2025

#### Contents


- Supplementary Methods

### 1 Supplementary Methods

#### 1.1 Examples for core notions

A criteria-satisfying symptom combination is called *irredundant* (for a certain disorder) if and only if no proper subset of the symptom combination is criteria-satisfying. Otherwise it is called *redundant* (for that disorder). In case of the disorder W in Fig. 2 the CSSC consisting of symptoms a,b,c is redundant since the presence of a,b is also a valid CSSC (and the latter is irredundant and so is the CSSC consisting of a,c).
We call a symptom *relevant* for diagnosing a certain disorder if and only if there exists an irredundant criteria-satisfying symptom combination containing it. Otherwise the symptom is called *irrelevant* (for diagnosing that disorder). In case of the disorder W in Fig. 2 each of the symptoms a,b,c are relevant, symptom d is irrelevant.
A CSSC is called *weakly irredundant* (for a certain disorder) if and only if it contains only relevant symptoms (or, equivalently, no irrelevant symptoms). Otherwise it is called *strongly redundant* (for that disorder). In case of the disorder W in Fig. 2 the CSSC consisting of symptoms a,b,c is weakly irredundant (as are a,b and a,c). The CSSC containing all of symptoms a,b,c,d, however, is strongly redundant.

The diagnostic criteria of two disorders (or, shortly, two disorders) are called *separable* if, and only if there does not exist any symptom combination that is weakly irredundant (and hence criteria-satisfying) for both disorders. Otherwise they are called *overlapping.* Clearly, overlapping diagnostic criteria do not allow to separate the corresponding disorders even if one considers only relevant symptoms (and hence excludes comorbidities). The disorders X and Y, e.g., considered in Fig. 2 are overlapping, because the CSSCs bc and bcd are weakly irredundant for both X and Y.
The diagnostic criteria of a disorder X are said to *subsume* the diagnostic criteria of a disorder Y if there exists a symptom combination that is weakly irredundant (and hence criteria-satisfying) for disorder X and also criteria-satisfying (but not necessarily weakly irredundant) for disorder Y. For example, in Fig. 2 disorder W subsumes disorder Z, because the symptom set a,b (as well as the symptom sets a,c and a,b,c) is weakly irredundant (and hence criteria-satisfying) for disorder W, but also criteria-satisfying for disorder Z due to the presence of symptom a (though strongly redundant for Z, because it also contains b, which is irrelevant for Z).
Note that subsumption is *not* a symmetric concept: X subsuming Y does (in general) *not* imply Y subsuming X. Moreover, subsumption is weaker than overlap: If the diagnostic criteria of two disorders X and Y overlap, then X subsumes Y and Y subsumes X, while the reverse need not be true. Hence subsumption is a weaker—and directed—form of overlap of the diagnostic criteria of two disorders.

#### 1.2 Implementation of the MPCS algorithm

Our implementation of MPCS uses a reversed row-column orientation of the matrix introduced in the main section. Adjusting the orientation of rows and columns reflects a more efficient algorithmic implementation, as many matrix operations benefit from having a larger number of rows than columns. Hence, the remaining paragraphs in this subsection use rows for the CSSCs and columns for the ordered symptoms.

We take advantage of Python's ability to efficiently handle matrix operations by computing all pairwise cosine similarities simultaneously. This is achieved through matrix-matrix multiplication combined with the outer product of the row-wise norms of both matrices. The outer product $A \otimes B$ of two vectors $A = (A_1, \ldots, A_n)$ and $B = (B_1, \ldots, B_m)$ is defined by

$$A \otimes B = AB^T = \begin{bmatrix} A_1 \\ A_2 \\ \vdots \\ A_n \end{bmatrix} [B_1, B_2, \ldots, B_m] = \begin{bmatrix} A_1B_1 & A_1B_2 & \cdots & A_1B_m \\ A_2B_1 & A_2B_2 & \cdots & A_2B_m \\ \vdots & \vdots & \ddots & \vdots \\ A_nB_1 & A_nB_2 & \cdots & A_nB_m \end{bmatrix} \quad (1)$$

where $B^T$ denotes the transpose of the vector $B$.

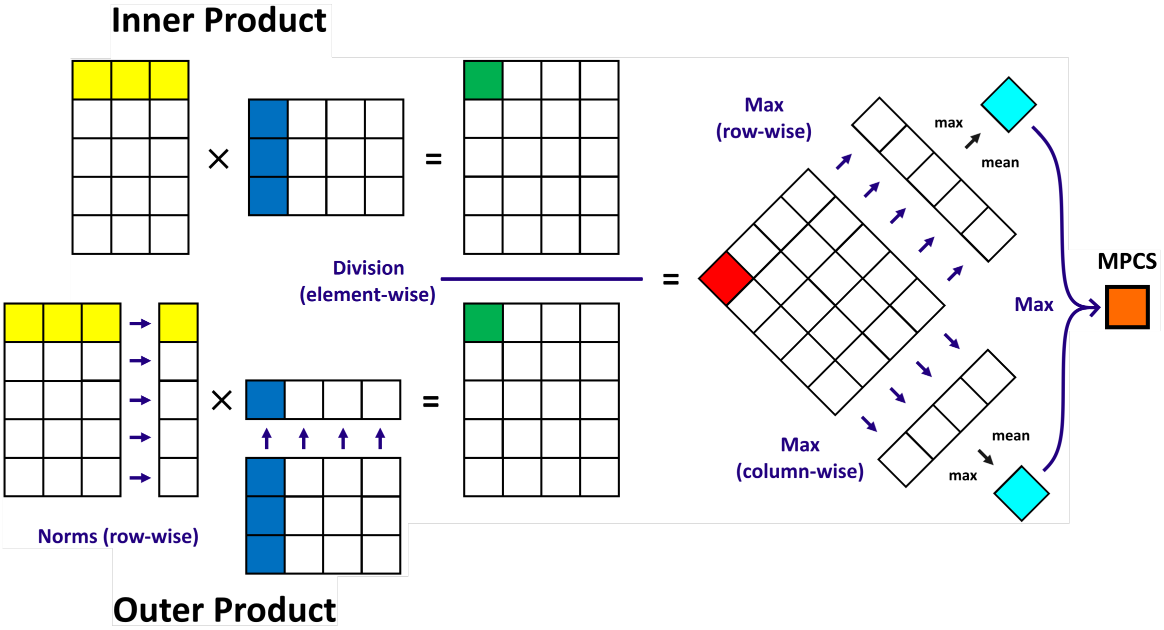


**Supplementary Figure 1:** Visualization of the MPCS algorithm implemented in Python.

The result is a similarity matrix with dimensions equal to the number of rows in matrix **B** by the number of rows in matrix **A**, enabling the element-wise (Hadamard) division between matrices to obtain the cosine similarity values. In other words: we obtain a matrix, in which every entry is the cosine similarity between a row vector from matrix **A** and one from matrix **B**. By taking the maximum of each column, we obtain the highest similarity for each row in **A**. Analogously, considering the maximum of each row yields the highest similarity for each row in **B**. Applying either the maximum or the mean aggregate to both of these lists, followed by calculating the maximum of the two values we finally obtain $\text{MPCS}_{\text{max}}$ and $\text{MPCS}_{\text{mean}}$ (see Supplementary Figure 1).

#### 1.3 Symptom harmonization examples

To align the Long COVID symptom definitions from the 2024 NASEM report with DSM-5 terminology, we performed a controlled mapping process. For example: "difficulty concentrating" was reformulated as "diminished ability to concentrate" (with "poor concentration" as a synonym); "lightheadedness" was supplemented with "feeling light-headed"; "hypersomnia" and "insomnia" were grouped under "sleep disturbances"; "fast heart rate" became "accelerated heart rate"; and gastrointestinal symptoms ("bloating," "constipation," "diarrhea") were aggregated under "abdominal distress." Only symptoms explicitly mentioned in the NASEM report were included.

To avoid diluting disorder-specific signal, we removed non-differentiating DSM-5 criteria such as "The episode is not attributable to the physiological effects of a substance or to another medical condition" (applies to all disorders), and "The symptoms cause clinically significant distress or impairment..." (excluded from MDD, PDD, and Generalized Anxiety Disorder. A descriptive clause in PDD—"Criteria for a major depressive disorder may be continuously present for two years"—was also excluded, as it is not a formal requirement.
